# Upregulation of TBX1 by genetic variants are associated with human congenital heart disease

**DOI:** 10.1101/2021.07.21.21260948

**Authors:** Liwei Yu, Binbin Li, Hongyan Wang

**Affiliations:** Children’s Hospital, Fudan University, Shanghai, China; Children’s hospital at Downstate, SUNY Downstate Medical Center, Brooklyn, NY, United States; Department of Molecular, Cellular, and Developmental Biology, University of Colorado Boulder, Boulder, CO, United States; Obstetrics and Gynecology Hospital, NHC Key Lab of Reproduction (Shanghai Institute of Planned Parenthood Research), Institute of Reproduction and Development, Fudan University, Shanghai, China; The Institutes of Metabolism and Integrative Biology, Fudan University, Shanghai, China

**Keywords:** TBX1, Congenital heart disease (CHD), Genetic variant, Single nucleotide polymorphism (SNP), Rare mutation

## Abstract

Congenital heart disease (CHD) is the most common human birth defect worldwide and also an important cause of childhood morbidity and mortality. The transcription factor of TBX1 early expressed in embryonic cardiac progenitor cells underlys embryo cardiogenesis in a dosage-dependent manner. Imbalanced TBX1 level has been shown to lead to cardiac defects. To study the association of *TBX1* genetic variants with CHD susceptibility, we screened genetic variants in 409 CHD patients and 203 healthy controls. One single nucleotide polymorphism (SNP), rs41260844, in TBX1 promotor region was identified to be associated with CHD. Functional studies showed the minor allele of rs41260844 is associated with higher CHD risk and increases *TBX1* promoter activity through attenuating *TBX1* promoter binding affinity with nuclear protein(s). In addition, a novel case-specific missense rare mutation of p.P164L in TBX1 T-box domain was identified and predicted as deleterious mutation, which showed a trend of increased protein function. In summary, we concluded that a higher TBX1 expression level or activity is associated with CHD susceptibility, which could affect TBX1 downstream targets and thus disrupt the balance of the complex regulation network during cardiogenesis. This study deepens our current understanding of embryo cardiogenesis and CHD etiology.

## Introduction

Congenital heart disease (CHD) is the most common type of birth defect in humans with the prevalence of around 4-10 per 1000 live births [1]. 1.35 million newborns are diagnosed with CHD each year [2]. Over the past couple decades, with multitude animal model studies and massive DNA sequencing conducted, numerous genes were suggested involved in cardiogenesis, among which transcription factors are one of the most important groups.

T-box family member, TBX1 marks the cardiac progenitor cells (CPCs) in secondary heart field (SHF), which can be considered as a reservoir of cardiac progenitors [3]. One important role for Tbx1 is to suppress differentiation and encourage proliferation of the CPCs [3, 4]. TBX1 facilitates proliferation of the CPCs in SHF before they migrate and differentiate to right ventricle myocardium, outflow tract (OFT), atrium myocardium and dorsal mesenchymal protrusion that are required for the right and left atrioventricular junctions [3, 5]. Once the progenitor cells move to OFT, TBX1 expression is no longer detectable [3], suggesting that its expression is highly spatially and temporally sensitive. *In vivo* study in *Tbx1* knock-out mouse embryos shows premature CPCs differentiation, and subsequent structural abnormality in outflow tract due to decreased number of cells [3]. Beyond that, TBX1 participates in the formation of the aortopulmonary septum, which divides aorta from pulmonary artery [4]. These findings are consistent with the cardiac deformities reported in mouse and human. Absence of TBX1 in mice leads to hypoplasia of distal OFT, common ventricular outlet, ventricular septal defect (VSD), and persistent truncus arteriosus (PTA) [6, 7]. And mutations in TBX1 have been reported in conditions including interrupted aortic arch (IAA), persistent truncus arteriosus (PTA), Tetralogy of Fallot (TOF), malaligned ventricular septal defect (VSD) [8].

Additionally, TBX1 is required for normal heart development in a dosage dependent manner [9]. *Tbx1*-null homozygous mouse embryo or newborns are not compatible for life secondary to the cardiac defects as mentioned above as well as craniofacial abnormalities such as shortened neck, low-set ears and abnormal facial structure, while *Tbx1* heterozygous mouse has milder phenotypes and lower penetrate rate [9-12]. On the other hand, mouse embryo with over-expressed TBX1 has similar phenotypes as DiGeorge Syndrome (22q11.2 deletion syndrome) and shows heart and thymic defect [13, 14]. In human, gain-of-function TBX1 mutation was reported to demonstrate DiGeorge like phenotype [15]. And the hallmark feature of 22q11.2 duplication syndrome is also OFT defects [16]. These results demonstrate that TBX1 has a dosage effect and needs to be tightly controlled to ensure appropriate cardiogenesis.

More than 30 genes were found to be TBX1 dependent and have a potential role in SHF [17]. They can be transcript factors, signaling pathway genes, muscle contraction genes, with either decreased, increased or even ectopic expression in *Tbx1* null mouse embryos [17]. *Pitx2* is believed to be one of the downstream genes regulated by TBX1, and they both are involved in asymmetric cardiac morphogenesis [18]. *Mef2c* is another downstream gene that is either directly and indirectly suppressed by TBX1 [19]. By suppressing Mef2c, muscle differentiation is suppressed [19], which is one major role of TBX1 in SHF [3, 17]. With regarding to the upstream regulation, TBX1 expression in SHF is upregulated by FOX proteins via Sonic Hedgehog pathway [20-22].

Given the dosage effect of this transcription factor, we designed this case-control study and screened genetic variants in both regulatory and coding regions of *TBX1* in human samples. Several novel SNPs and non-synonymous rare mutations were identified. One SNP, rs41260844, was identified associated with CHD occurrence, the minor allele of which increases *TBX1* promoter activity and is accumulated in CHD patients. The novel case-specific missense rare mutation of p.P164L in T-box domain of TBX1 is suggested as deleterious mutation *in Silicon*. Hence, we conclude that harmful genetic variants leading to imbalanced TBX1 activity is strongly associated with CHD in humans.

## Result

### Minor allele of rs41298814 is a risk factor of CHD occurrence

409 CHD patients and 213 healthy controls were recruited for genetic variant screening. Nine SNPs with minor allele frequency (MAF) > 5% in *TBX1* regulatory and coding regions were identified (Table 1). The association study showed that the distribution of major and minor allele of rs41260844 (T and C, respectively) is significantly different between case and control. The minor allele of rs41260844 is significantly enriched in cases (*p* = 0.0041, OR = 1.42, 95% CI = 1.11-1.81), which means that this minor allele is associated with higher risk of CHD (Table 1).

**Table 1.**
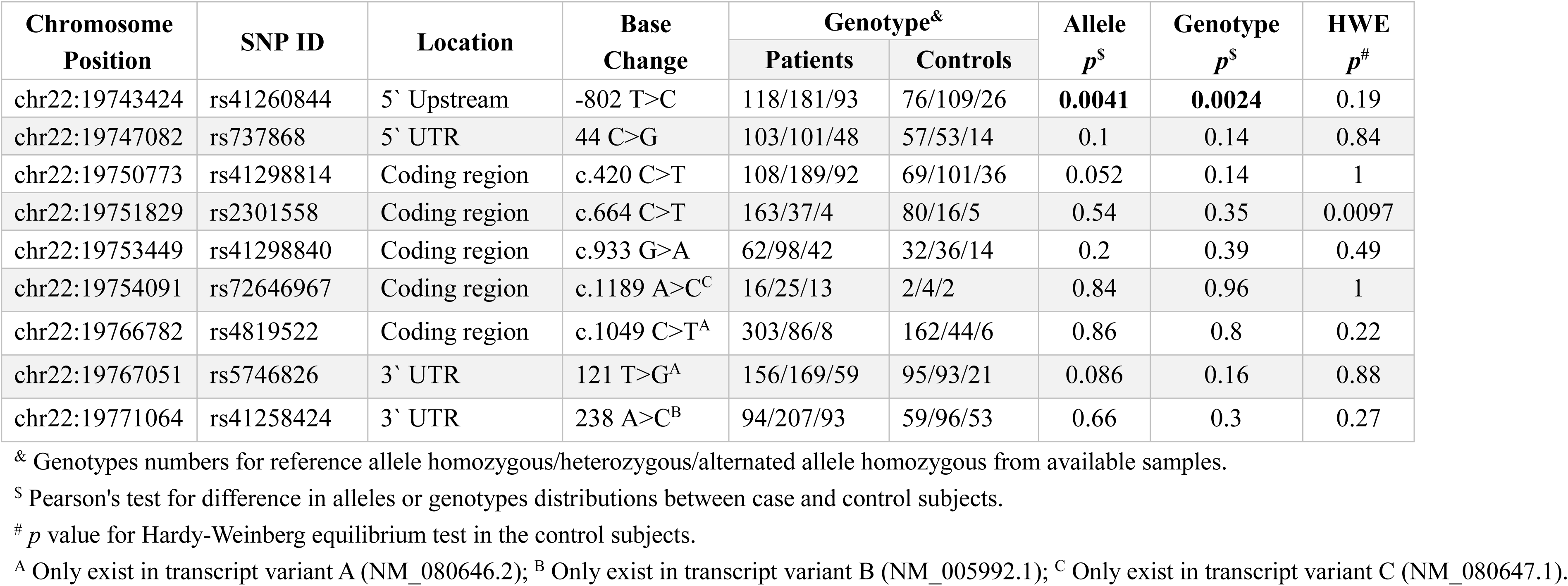
Summary of *TBX1* common variants.

We then analyzed all 9 SNP in haplotypes. Three out of nine SNPs, rs41298814, rs41298840 and rs72646967, were in linkage disequilibrium (R^2^> 0.8), (Fig S1). Given that rs41298814 is the SNP that shows association with CHD, it was listed as tag SNP and analyzed with other 6 SNPs for haplotype. A total of 15 haplotypes were identified in our sample, and one haplotype is associated with increased risk of CHD (*p*= 0.0049, OR= 2.38, 95% CI=1.01-5.65) (Table S1). It contains minor allele of rs41298814, which is also a risk factor of CHD in the individual SNP association study above.

### Minor allele of rs41260844 upregulates *TBX1* expression and attenuates *TBX1* promoter binding affinity with nuclear proteins

To test the potential role of different genotype of rs41260844 on regulating *TBX1* promoter activity, a *TBX1* promoter fragment containing major or minor allele of rs41260844 was amplified and subcloned into pGL3-Basic reporter to drive firefly luciferase expression (termed as Major (T) and Minor (C), respectively), and subsequently tested in HEK 293T cells as well as the rat cardiomyocyte derived H9C2 cells. In both cell lines, both major and minor allele containing promotor could activate luciferase expression robustly, compared with empty vector group. Remarkably, the minor allele containing promoter exhibits significantly higher luciferase expression than that of major allele (Fig. 1). Given that minor allele is associated with higher CHD risk, we conclude that higher TBX1 level is associated with higher CHD risk.

**Fig 1.**
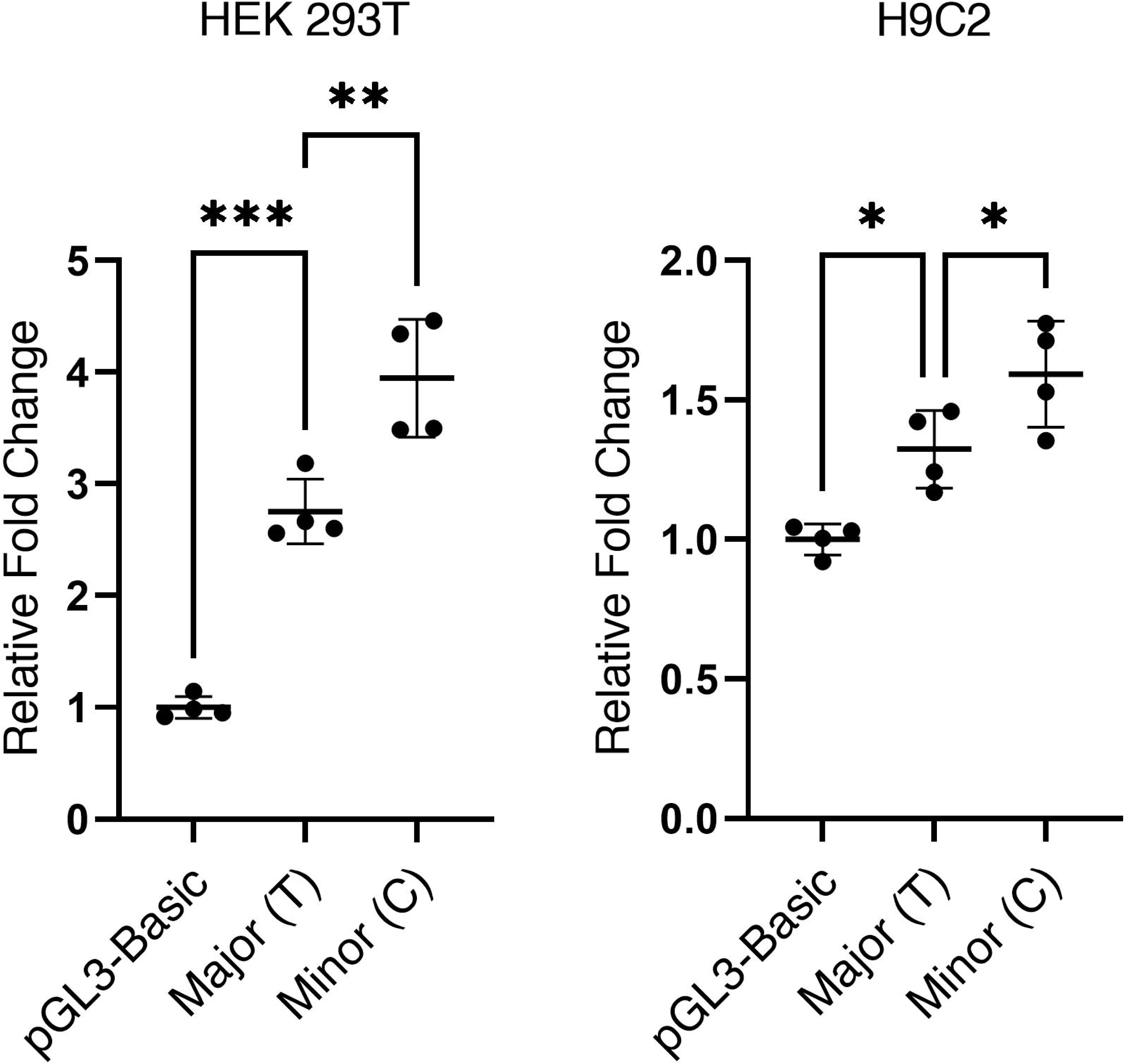
Dual luciferase assay to analyze different allele of rs41260844. Luciferase assay of *TBX1* promoter activity were performed in HEK 293T cells (A) and H9C2 cells (B). Constitutively expressed Renilla reniformis luciferase served as an internal control. (n=4; \**p*<0.05; \*\**p*< 0.01; \*\*\**p*<0.001, compared with Major (T) allele group).

In order to test if the change in *TBX1* promotor activity is due to different affinity with nuclear proteins, we then performed Electrophoretic mobility shift assay (EMSA) using nuclear extract of H9C2 cells and DNA probes in 2 genotypes. Probe-major has T allele of rs41260844 and probe-minor has C allele of rs41260844. Both of them are synthesized in 2 forms: biotin labelled probe and un-labelled probe. The result shows a specific band pointed by arrow, which has lower affinity with minor probe (Fig. 3). This study showed that the level of promotor activity is probably influenced by altered binding affinity with certain transcriptional factor. Given that *TBX1* promotor with minor allele has higher transcription activity but lower binding affinity with nuclear protein, it is possible that the minor allele of rs41260844 attenuates *TBX1* promoter binding affinity with a negative regulator.

**Fig 2.**
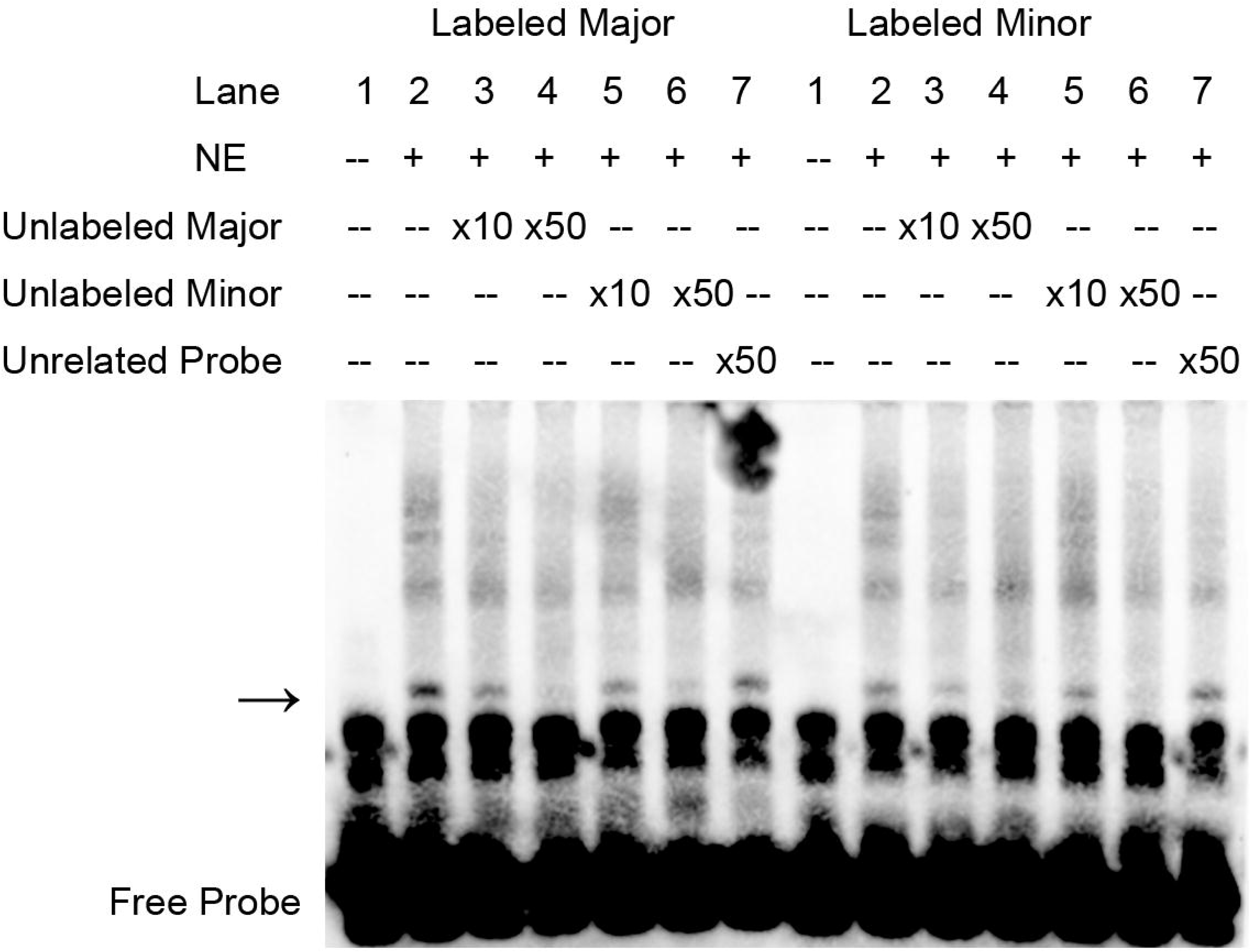
Analysis of DNA binding affinity with two genotypes. Binding affinity of Major/Minor-probes with H9C2 nuclear proteins. Highlighted arrow pointed out one specific nuclear protein. Minor probe has weaker binding affinity.

**Fig 3.**
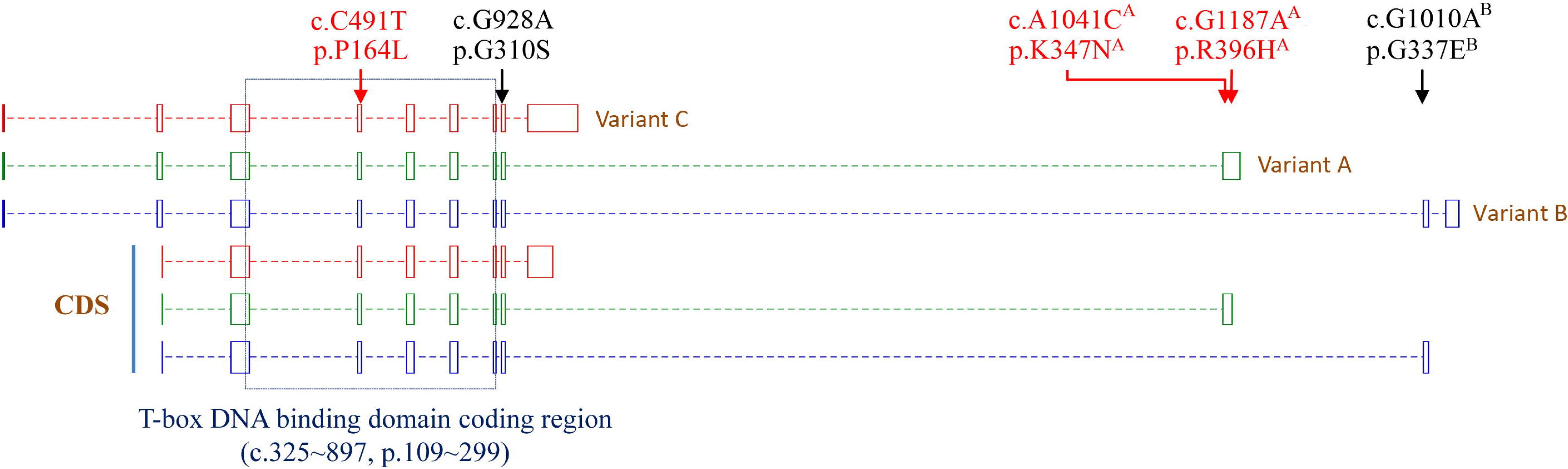
TBX1 non-synonymous rare mutations identified in CHD patients. Rare mutations are mapped in TBX1 transcript variant A (NM_080646.2), variant B (NM_005992.1) and variant C(NM_080647.1). Variant A, B and C have same translation starting site. Variant D (NM_001379200.1) with a distinguished translation starting site is not displayed and there is no variant D only mutation. Mutations in red is case specific, mutations in black are also found in controls or databases.

### Identification of rare variants in *TBX1* gene of CHD patients

Rare genetic variants (MAF< 5%) in non-coding region are listed in Table S2, which are considered less important. In coding region, 8 rare mutations were identified (Table 2 and Fig 3). The three misssense rare mutations, p.P164L, p.K347N and p.R396H are case-specific mutations. p.K347N and p.R396H only exist the protein isoform from transcript variant A while p.P164L locates on T-box binding domain of a highly conserved structure in all TBX1 isoforms, indicating that this mutation probably affects TBX1 binding affinity with target DNA motif. Indeed, both broadly accepted bioinformatics tools, SIFT and Polyphen-2, strongly suggest that p.P164L is a harmful mutation (Table 2). Further functional study is desired to validate the negative effect caused by this novel rare mutation.

**Table 2.** Summary of rare mutations in *TBX1* coding region.

| Chromosome<br>Position | SNP ID | Location | Base change | Function | Genotype<sup>&</sup> |  | SIFT<sup>*</sup> | PolyPhen-2<sup>#</sup><br>(HumDiv) |
| --- | --- | --- | --- | --- | --- | --- | --- | --- |
|  |  |  |  |  | Patients | Controls |  |  |
| **Chr22:19750815** | rs41298816 | Coding region | c.462 C>T | Synonymous | 390/1/0 | 205/0/0 | - | - |
| **Chr22:19750844** | - | Coding region | c.491 C>T | p.P164L | 387/1/0 | 202/0/0 | 0 | 1 |
| **Chr22:19753444** | rs41298838 | Coding region | c.928 G>A | p.G310S | 183/14/2 | 78/4/0 | 0.57 | 0.835 |
| **Chr22:19754066** | - | Coding region | c.1164 G>C<sup>C</sup> | Synonymous | 16/1/0 | 2/0/0 | - | - |
| **Chr22:19766774** | - | Coding region | c.1041 A>C<sup>A</sup> | p.K347N | 395/1/0 | 212/0/0 | 0.34 | 0.01 |
| **Chr22:19766918** | - | Coding region | c.1185 C>T<sup>A</sup> | Synonymous | 397/0/0 | 208/4/0 | - | - |
| **Chr22:19766920** | - | Coding region | c.1187 G>A<sup>A</sup> | p.R396H | 396/1/0 | 212/0/0 | 0.1 | 0.018 |
| **Chr22:19770436** | rs180794829 | Coding region | c.1010 G>A<sup>B</sup> | p.G337E | 394/1/0 | 206/0/0 | 1 | 0.008 |

<sup>A</sup> Only exist in transcript variant A (NM\_080646.2); <sup>B</sup> Only exist in transcript variant B (NM\_005992.1); <sup>C</sup> Only exist in transcript variant C (NM\_080647.1).
Mutations without superscript exist in all variants.
<sup>&</sup> Genotypes numbers for reference allele homozygous/heterozygous/alternated allele homozygous from available samples.
<sup>\*</sup> Deleterious threshold for SIFT: < 0.05; <sup>#</sup> Deleterious threshold for PolyPhen-2: > 0.5.

## Discussion

*TBX1* is the most import gene absent in DiGeorge syndrome [23]. This correlation of phenotype and genotype is not commonly seen in other transcriptional factors that are important to cardiogenesis, which makes TBX1 a good candidate to understand the genetic etiology of CHD. Most of the studies on human genetic variants identified in CHD are loss of function [24-26]. Given the dosage effect of TBX1, we hypothesis that genetic variants that upregulate TBX1 transcription or activity may also exist in CHD patients. Current case-control study demonstrated that the minor allele of rs41260844 enriched in CHD patients increases *TBX1* promotor activity. Given the irreplaceable role of TBX1 played in SHF during cardiogenesis as illustrated in Introduction, our study provides new evidence showing redundant TBX1 is also harmful to embryo heart morphogenesis.

There are other two studies on rs41260844 [27, 28]. One of them was aimed at a different disease, inguinal hernia, while no association between variant and disease was found, and no change of *TBX1* promotor activity between two genotypes in human lung fibroblast cell (HLF-1) [28]. The other one was from the same institute but focused on congenital heart disease [27], in which they identified the SNP by comparing allele frequencies of ventricular septal defect (VSD) patients and healthy controls. Interestingly, their study revealed the same result as we do: minor allele of rs41260844 is more frequent in CHD patients and generated higher promotor activity in HEK 293T cells. TBX1 is expressed in outflow tract and myocardium of atrium and ventricle [5], which makes sense that more types of congenital heart disease can be related to *TBX1* genetic variants. Our case samples are not only from VSD, and the majority of our samples consist of outflow tract defect and septal defects (Table S3). In addition, our study also tested TBX1 promotor activity in cardiomyocyte derived H9C2 cells and got the same trend of result (Fig. 1). Thus, our result more accurately reflects how this genetic variant could possibly act like in CHD.

Our EMSA result shows there is a potential nuclear protein specifically binding TBX1 promoter and the minor allele attenuates this binding affinity in cardiomyocyte cells (Fig. 2). Given that minor allele encourages a higher promotor activity as we and the other group demonstrated, the possible nuclear protein could be a negative regulator. However, we had a hard time finding the possible upstream proteins that could negatively regulate TBX1 expression. So far, no known negative regulator of TBX1 has been reported. More studies are needed to identify exact nuclear protein(s) that with binding motif consisting of rs41260844.

After we further looked into the data, the female patient who carries p.P164L rare mutation has Tetralogy of Fallot (TOF). The rare mutation is predicted as harmful, and was found as a gain of function mutation in our preliminary experiment in which we co-transfected TBX1 wild type or mutant expression plasmid with a homemade luciferase reporter driven by TBX1 binding motif containing promoter in HEK 293T cells[29]. We were unable to achieve robust reporter activation, but we saw slight increasement in mutant group. (Data not shown). Developing an optimized TBX1 responsive reporter is deserved in our future work. It worth noting that the patient who possesses this mutation also carries homozygous minor allele of rs41260844 (C/C), which increases *TBX1* transcription as demonstrated above. Her CHD type matches with the potential phenotype from altered TBX1 function [15, 23]. Thus, there is a possibility that both the harmful rare mutation and the minor alleles of the SNP coordinately elevated the impact of TBX1 in the regulatory network, and eventually made the individual more susceptible to CHD than others carrying no or one type of harmful variant. This finding fits the previous report of our group that accumulation of multiple harmful genetic variants in single individual could probably add to the approaching the threshold of complex reproductive disadvantage disease like CHD [30].

To summarize, we identified both common and rare genetic variants of *TBX1* gene which lead to dysregulated TBX1 transcription/activity and are associated with CHD occurrence. Our result broadens the current knowledge that precise TBX1 regulation is critical for normal embryo cardiogenesis. This also enhances our understanding of the genomic architecture underlying susceptibility to this common congenital malformation of CHD.

## EXPERIMENTAL PROCEDURES

### Study subjects and identification of genetic variants

All CHD patients (n=409, 3±2.7 years, 54.6% males) were recruited from Cardiovascular Disease Institute of Jinan Military Command (Jinan, Shandong, China). The diagnosis was made via echocardiography, some were further confirmed with surgery. Patients with clinical features of syndromic diseases, positive family history of CHD in a first-degree relative, maternal diabetes mellitus, maternal exposure to known teratogens or any therapeutic drugs during gestation were excluded. For detailed classification of CHD subtypes, please refer to Supplementary Table S3. Blood samples from the 213 ethnically and gender-matched unrelated healthy controls (7.1±3.7 years, 49.8% males) were also recruited from the same region, and individuals with any congenital anomalies or cardiac disease were excluded. All samples were collected with the approval of local ethics committees and institutional review board of Fudan University. Informed consent documents were signed by the parents or guardians.

Genomic DNA was extracted and targeted exome pooling sequencing of 258 candidate genes including *TBX1* was conducted as described previously [31]. Identified variants were filtered using the dbSNP database (https://www.ncbi.nlm.nih.gov/projects/SNP/) and the 1000 genomes project (http://www.1000genomes.org). All the case-specific non-synonymous mutations were subsequently confirmed by Sanger DNA sequencing.

### Plasmid construction, cell culture, and luciferase assays

To construct luciferase reporter plasmids with the *TBX1* promoter, we amplified 1,855 bp fragment of *TBX1* c.-1,797 to c.64 (GRCh38.p13, Chr22: 19754912-19756766) directly from samples carrying homozygous major allele (T/T) or minor allele (C/C) of rs41260844 and subcloned into KpnI/XhoI sites of pGL3-Basic plasmid (Promega, USA) and named as Major (T) and Minor (C), respectively.

HEK 293T cells and H9C2 cells were obtained from the American Type Culture Collection (ATCC, #CRL-3216 and # CRL-1446, respectively) and cultured in high-glucose Dulbecco’s Modified Eagle Medium (DMEM, Thermo Fisher Scientific, #11995065) supplemented with 10% FBS (Thermo Fisher Scientific, #A3840001) at 37_°C with 5% CO2. Cells were seeded and maintained overnight to reach 80% confluency at the time of transfection. Lipofectamine 3000 Transfection Reagent (Thermo Fisher Scientific, #L3000015) was used for plasmid transient transfection, following manufacturer’s protocol.

### Dual luciferase reporter assay

Reporter assays were performed as described previously [32, 33]. Briefly, each well of 24-well plate was co-transfected with 300ng of pGl3-Basicfirefly luciferase empty vector or Major (T) or Minor (C), and 10ng of Renilla reniformis luciferase reporter (pGL4.74[hRluc/TK], Promega, #E6921) serving as an internal control. Reporter assays were performed 24 h post transfection using the Dual-Luciferase Reporter Assay System (Promega, #E910) on GloMax Navigator Microplate Luminometer (Promega, #GM2010).

### Probe design and Electrophoretic mobility shift assay (EMSA)

Based on the two genotypes of rs41260844, two probes labeled with biotin were named as Labeled Major and Labeled Minor, respectively. The other two Unlabeled Major and Unlabeled Minor, as well as an unlabeled unrelated probe were also synthesized for competing assay.

Nuclear extracts were prepared from H9C2 cells using the NE-PER Nuclear and Cytoplasmic Extraction kit (Life Technologies, USA) and stored at -80 °C before use. Protein concentration was measured using Pierce BCA Protein Assay Kit (ThermoFisher Scientific, USA#23227). Nuclear proteins protein was mixed with indicated probe and incubated, and subsequently processed using LightShift Chemiluminescent EMSA Kit (ThermoFisher Scientific, #20148) following manufacturer’s instruction.

### Statistical analysis

Hardy–Weinberg equilibrium in the controls and the association study in both the CHD cases and the controls were compared using the χ2 test. We calculated the odds ratios (ORs) and 95% confidence intervals (CIs). The association study between SNPs or haplotypes and CHD risk were performed using the SNPStats (https://www.snpstats.net/start.htm). We reconstructed haplotypes of the SNPs using Haploview version 4.2.

Differences in cell culture study were evaluated using the Oneway ANOVA. The Oneway ANOVA was two-tailed with p<0.05 as the significance level and were performed using the SPSS 16.0 software (SPSS, USA).

## Data Availability

The original contributions presented in the study are included in the article/Supplementary Material, further inquiries can be directed to the corresponding author.

## Author contribution

LY, BL, HW and YG designed the project. LY and BL performed the experiments. LY, BL, HW and YG prepared the manuscript.

## Acknowledgement

We sincerely thank all the subjects who participated in this study and the clinicians who helped in recruiting the CHD cases and controls.

## Supplementary information

**Table S1.**
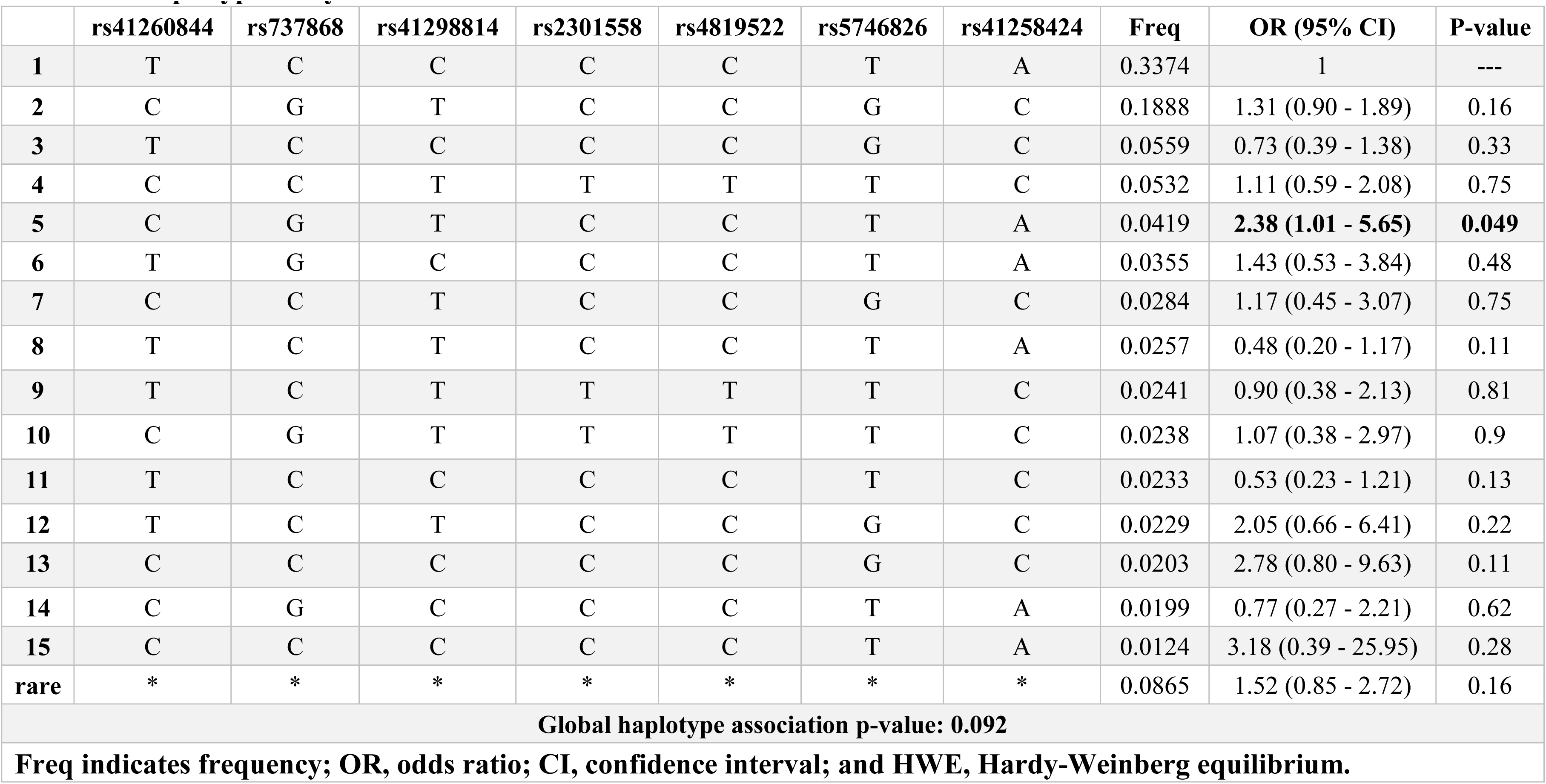
Haplotype analysis of *TBX1* common variants.

**Table S2.** Summary of rare mutations in *TBX1* non-coding region.

| Chromosome<br>Position | SNP ID | Location | Base change | Genotype<sup>&</sup> |  |
| --- | --- | --- | --- | --- | --- |
|  |  |  |  | Patients | Controls |
| **Chr22:19742426** | rs112910011 | 5` Upstream | C>T | 395/0/0 | 210/1/0 |
| **Chr22:19742522** | rs41298615 | 5` Upstream | C>T | 390/3/0 | 208/0/0 |
| **Chr22:19742546** |  | 5` Upstream | G>C | 390/1/0 | 210/0/0 |
| **Chr22:19743061** |  | 5` Upstream | G>A | 212/4/0 | 105/0/0 |
| **Chr22:19743185** |  | 5` Upstream | C>G | 343/2/0 | 180/0/0 |
| **Chr22:19743205** |  | 5` Upstream | C>T | 358/0/0 | 191/1/0 |
| **Chr22:19743400** |  | 5` Upstream | T>G | 393/1/0 | 210/0/0 |
| **Chr22:19743473** | rs41298629 | 5` Upstream | C>T | 387/6/0 | 208/1/0 |
| **Chr22:19743530** |  | 5` Upstream | C>T | 389/0/0 | 203/1/0 |
| **Chr22:19743735** |  | 5` Upstream | A>C | 369/0/0 | 199/2/0 |
| **Chr22:19743806** |  | 5` Upstream | T>C | 396/0/0 | 206/1/0 |
| **Chr22:19743963** | rs149780269 | 5` Upstream | C>T | 398/1/0 | 210/0/0 |
| **Chr22:19744029** | rs112229231 | 5` Upstream | C>T | 400/0/0 | 210/1/0 |
| **Chr22:19744078** |  | 5` Upstream | C>T | 399/0/0 | 208/1/0 |
| **Chr22:19747128** | rs72646950 | 5` UTR | C>T | 317/0/0 | 150/1/0 |
| **Chr22:19753213** |  | Intronic | C>T | 381/1/0 | 199/1/0 |
| **Chr22:19753357** |  | Intronic | C>A | 341/2/0 | 171/0/0 |
| **Chr22:19753374** | rs199883948 | Intronic | A>C | 309/4/0 | 148/1/0 |
| **Chr22:19754403** |  | 3` UTR | C>T | 320/2/0 | 152/0/0 |
| **Chr22:19754720** |  | 3` UTR | C>T | 367/1/0 | 192/0/0 |
| **Chr22:19754804** |  | 3` UTR | A>G | 353/2/0 | 184/0/0 |
| **Chr22:19766954** | rs41298008 | 3` UTR | C>G | 386/9/0 | 209/4/0 |
| **Chr22:19770886** | rs9680615 | 3` UTR | G>A | 370/25/1 | 193/16/1 |
| **Chr22:19770973** | rs41298852 | 3` UTR | T>C | 390/8/0 | 207/2/0 |
| **Chr22:19771054** |  | 3` UTR | C>T | 392/2/0 | 209/0/0 |

<sup>&</sup> Genotypes Numbers for reference allele homozygous/heterozygous/alternated allele homozygous from available samples.

**Table S3.**
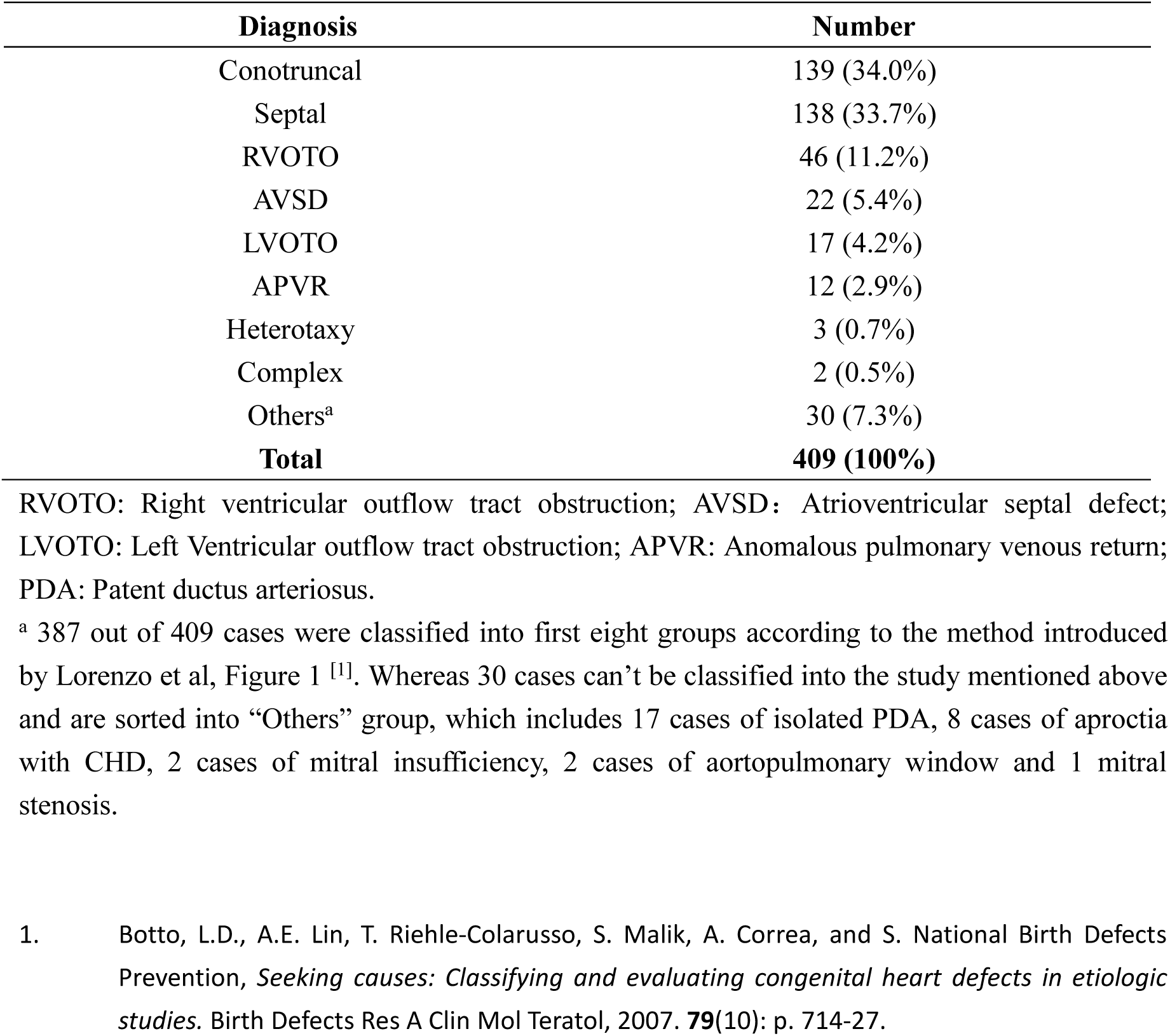
Classification of CHD subtypes in our study.

